# Admixture Mapping of Peripheral Artery Disease in a Dominican Population Reveals a Novel Risk Locus on 2q35

**DOI:** 10.1101/2023.03.27.23287788

**Authors:** Sinead Cullina, Genevieve L. Wojcik, Ruhollah Shemirani, Derek Klarin, Bryan R. Gorman, Elena P. Sorokin, Christopher R. Gignoux, Gillian M. Belbin, Saiju Pyarajan, Samira Asgari, Phil S. Tsao, Scott M. Damrauer, Noura S. Abul-Husn, Eimear E. Kenny

**Affiliations:** Institute for Genomic Health, Icahn School of Medicine at Mount Sinai, New York, NY; Department of Genetics and Genomic Sciences, Icahn School of Medicine at Mount Sinai, New York, NY, USA; Department of Epidemiology, Johns Hopkins Bloomberg School of Public Health, Baltimore, MD, USA; Division of Genomic Medicine, Department of Medicine, Icahn School of Medicine at Mount Sinai, New York, NY, USA; Department of Genetics, Stanford University, Stanford, California, CA; Division of General Internal Medicine, Department of Medicine, Icahn School of Medicine At Mount Sinai, New York, NY, USA; VA Palo Alto Healthcare System, Palo Alto, CA USA; Division of Vascular Surgery, Stanford University School of Medicine, Palo Alto, CA, USA; Center for Data and Computational Sciences (C-DACS), VA Boston Healthcare System, Boston, MA, USA; Booz Allen Hamilton, McLean, VA, USA; Calico Life Sciences LLC, South San Francisco, USA; Human Medical Genetics and Genomics Program, University of Colorado Anschutz Medical Campus, Aurora, CO, USA; Department of Biomedical Informatics, University of Colorado Anschutz Medical Campus, Aurora, CO, USA; Colorado Center for Personalized Medicine, Aurora, CO, USA; Department of Medicine, Brigham Women’s Hospital, Harvard Medical School, Boston, MA, USA; Corporal Michael J. Crescenz VA Medical Center, Philadelphia, PA, USA; Department of Surgery, Perelman School of Medicine, University of Pennsylvania, Philadelphia, PA, USA; 23andMe, Sunnyvale, CA, USA; Department of Genetics, University of Pennsylvania Perelman School of Medicine, Philadelphia, PA, USA

## Abstract

Peripheral artery disease (PAD) is a form of atherosclerotic cardiovascular disease, affecting ∼8 million Americans, and is known to have racial and ethnic disparities. PAD has been reported to have significantly higher prevalence in African Americans (AAs) compared to non-Hispanic European Americans (EAs). Hispanic/Latinos (HLs) have been reported to have lower or similar rates of PAD compared to EAs, despite having a paradoxically high burden of PAD risk factors, however recent work suggests prevalence may differ between sub-groups. Here we examined a large cohort of diverse adults in the Bio*Me* biobank in New York City (NYC). We observed the prevalence of PAD at 1.7% in EAs vs 8.5% and 9.4% in AAs and HLs, respectively; and among HL sub-groups, at 11.4% and 11.5% in Puerto Rican and Dominican populations, respectively. Follow-up analysis that adjusted for common risk factors demonstrated that Dominicans had the highest increased risk for PAD relative to EAs (OR=3.15 (95% CI 2.33-4.25), *P*<6.44×10^-14^). To investigate whether genetic factors may explain this increased risk, we performed admixture mapping by testing the association between local ancestry (LA) and PAD in Dominican Bio*Me* participants (N=1,940) separately for European (EUR), African (AFR) and Native American (NAT) continental ancestry tracts. We identified a NAT ancestry tract at chromosome 2q35 that was significantly associated with PAD (OR=2.05 (95% CI 1.51-2.78), *P*<4.06×10^-6^) with 22.5% vs 12.5% PAD prevalence in heterozygous NAT tract carriers versus non-carriers, respectively. Fine-mapping at this locus implicated tag SNP rs78529201 located within a long intergenic non-coding RNA (lincRNA) *LINC00607*, a gene expression regulator of key genes related to thrombosis and extracellular remodeling of endothelial cells, suggesting a putative link of the 2q35 locus to PAD etiology. In summary, we showed how leveraging health systems data helped understand nuances of PAD risk across HL sub-groups and admixture mapping approaches elucidated a novel risk locus in a Dominican population.

## Introduction

Peripheral arterial disease (PAD) is a form of atherosclerotic disease leading to peripheral artery obstruction. PAD is characterized by classic symptoms of intermittent claudication of the lower extremities, and ankle-brachial systolic pressure index (ABI) <0.9 (Crawford et al., 2016). PAD is a common disease affecting ∼8 million Americans (Leeper et al., 2012) with a combined annual health care cost exceeding $21 billion in the United States (US) (Mahoney et al., 2008). Symptomatic PAD negatively impacts quality of life for patients, and severe outcomes include chronic ischemia and amputation (Elnady and Saeed, 2017; Klarin et al., 2021). Even when asymptomatic, PAD is associated with systemic vascular disease and increased rates of myocardial infarction, stroke and death (American Diabetes Association, 2003). The incidence of PAD at a population level depends on a nuanced interplay between ancestral, social, and environmental factors (Allison et al., 2010; Wassel et al., 2011; Klarin et al., 2021). Non-genetic risk factors include diabetes, smoking, thrombosis, increased age, systolic blood pressure, C-reactive protein levels, and serum total cholesterol (Kannel and McGee, 1985; Wassel et al., 2011; Wu et al., 2019). Smoking and thrombosis, in particular, are thought to have a larger contribution to PAD etiology compared to other circulatory diseases (Klarin et al., 2021). The heritability of PAD based on family studies is estimated to be 20% to 30% (Klarin et al., 2021). Genome-wide association studies (GWAS) of PAD have provided further insights into the biological pathways contributing to this polygenic disease (Thorgeirsson et al., 2008; Kullo et al., 2014; Matsukura et al., 2015). The largest PAD GWAS conducted to date is in a multi-ancestry Million Veteran Program (MVP) cohort which identified 19 genome wide significant (GWS) loci. Genes underlying PAD associated loci are associated with biological processes linked to known PAD risk factors, including type 2 diabetes, smoking habits, hypertension, lipids and thrombosis (Klarin et al., 2019, 2021).

There are known racial and/or ethnic disparities in incidence rates of PAD (Allison et al., 2007; Shu and Santulli, 2018; Belbin et al., 2021; Hackler et al., 2021). African Americans (AAs) are reported to have the highest rates of PAD as well as PAD associated amputation compared to other race and/or ethnicity groups in the US (Shu and Santulli, 2018; Matsushita et al., 2019; Hackler et al., 2021). Studies have suggested that rates of PAD in Hispanic/Latinos (HLs) are lower than both non-Hispanic Whites and AAs (Allison et al., 2006), however there is also evidence that PAD rates may differ across HL sub-groups, with higher rates in HL groups with origins in the Caribbean (Daviglus et al., 2012; Allison et al., 2015). Previous phenome-wide association analysis from our group in the diverse Bio*Me* biobank in a large health system in New York City (NYC), observed enrichment of PAD in both AAs, and in two HL sub-groups, Puerto Rican and Dominican. Of the three populations, the strongest enrichment was observed in the Dominican population (Belbin et al., 2021).

Admixture mapping is a powerful approach for genomic discovery when it is suspected that prevalence of disease and/or frequency of underlying causal variants may differ across populations. Unlike GWAS approaches, which treat population structure as a confounder, admixture mapping leverages genetic ancestry differences at a haplotypic level, usually in populations with recent ancestry from two or more continents, to test for correlation with health outcomes of interest (Winkler et al., 2010). Examples of admixture mapping discoveries include an association of *APOL1* and renal disease in AAs (Kao et al., 2008; Kopp et al., 2008), and numerous loci with Alzheimer’s disease in HLs (Horimoto et al., 2021; Kizil et al., 2022). Notably, admixture mapping previously identified rs9665943 as being a risk locus for PAD (ankle-arm index) in AAs (Scherer et al., 2010).

Here we examined the prevalence of PAD in self-reported race and/or ethnicity groups, and in sub-groups inferred using genetic ancestry, in the Bio*Me* biobank at the Mount Sinai health system in NYC. We demonstrate that prevalence of PAD is similarly high in self-reported HLs and AAs, and within sub-groups, Dominicans have the highest PAD risk, even after adjusting for age, sex, body mass index, type 2 diabetes, triglycerides, total cholesterol, and high-density lipoprotein. We performed genomic discovery analysis using admixture mapping approaches in the Dominican population to test whether any regions were associated with increased risk for PAD. Admixture mapping discovered significant association between Native American genetic ancestry haplotypes and PAD on 2q35. Fine-mapping at this locus implicated a tag SNP rs78529201 located within long intergenic non-coding RNA (lincRNA) *LINC00607*. Existing literature highlights *LINC00607* as a super enhancer in a gene expression network that is critical for regulation of normal endothelial cell function as well as dysfunction, suggesting a putative link of the 2q35 locus to PAD etiology (Calandrelli et al., 2019; Boos et al., 2022; Sriram et al., 2022).

## Materials and methods

### Study population

The Bio*Me* biobank is an electronic health record-linked biorepository that has been enrolling participants from across the Mount Sinai health system in NYC since 2007. There are currently over 50,000 participants enrolled in the Bio*Me* biobank under an Institutional Review Board (IRB)-approved study protocol and consent (IRB 07–0529). Recruitment occurs predominantly through ambulatory care practices, and participants consent to provide whole blood derived germline DNA and plasma samples which are banked for future research. Participants also complete a questionnaire providing personal and family history as well demographic and lifestyle information as has been previously described (Belbin et al., 2021) and (Abul-Husn et al., 2021). Bio*Me* participants represent the broad diversity of the New York metropolitan area; and greater than 65% of participants represent minority populations in the US. All participants provided informed consent, and the study was approved by the Icahn School of Medicine at Mount Sinai’s IRB (protocol number 07-0529).

### Using self-reported and genetic ancestry information to define population groups

All participants were asked multiple choice questions at enrollment regarding their heritage and country-of-birth of self, parents, and grandparents, which have been previously described in (Belbin et al., 2021). Participants’ responses to the heritage question were mapped into eight single self-report groups for this study; namely AA, East/South-East Asian (EAsn), South Asian (SA), Native American (NA), EA, HL, Other and Multiple Selected. Those participants who selected either ‘Hispanic/Latino’ alone, or ‘Hispanic/Latino’ in addition to one or more other categories, were designated as HL. Participants who selected ‘White/Caucasian’ and/or ‘Ashkenazi Jewish’, were designated as EA. Participants who selected either ‘Mediterranean’ or ‘Other’ or a combination of both were assigned Other, and those with any other combination of multiple race ethnicity labels were assigned Multiple Selected.

Genetically inferred ancestry information was used to designate sub-populations. In brief, array data (Illumina omni-express (OMNI) and Illumina multi-ethnic global array (MEGA)) was phased and used to infer pairwise shared haplotypes identical by descent (IBD). Unsupervised clustering methods based on population-level IBD sharing were used to define clusters or IBD communities of individuals sharing recent cryptic relatedness. Over 50% of participants provided responses to questions about their country of birth which were used to determine the positive predictive value (PPV) of IBD communities in detecting recent patterns of diaspora to NYC. For example, one IBD community (N=2075) had a high confidence of predicting (PPV = >0.9) individuals who were born, or who had parents or grandparents born, in the Dominican Republic and was designated the Dominican community; PPVs and definitions of all IBD communities used in downstream analysis are described in detail in Belbin *et al (Belbin et al., 2021)*, and reported in **Table 1**.

**Table 1:** Characteristics of BioMe participants showing PAD prevalence across PAD risk factors, self-reported and IBD community membership. P-values represent a chi-squared test for discrete variables and Mann-Whitney test for continuous variables.

| Characteristics of the Study Participants |  |  |  |  |
| --- | --- | --- | --- | --- |
|  | N | PAD cases | PAD controls | P value |
| **All BioMe participants (N, %)** | 57580 | 3762 (6.5%) | 53818 (93.5%) |  |
| Sex (N women, %) | 57776 | 2049 (54.4%) | 31422 (58.2%) | $6.95 \times 10^{-06}$ |
| Age (median years, IQR) | 57671 | 73 (16) | 59 (28) | $2.22 \times 10^{-308}$ |
| BMI (median kg/h<sup>2</sup>, IQR) | 59340 | 28.2 (8.2) | 26.7 (7.8) | $1.60 \times 10^{-51}$ |
| T2D (N cases, %) | 51532 | 2130 (66.9%) | 9327 (19.2%) | $2.22 \times 10^{-308}$ |
| Total Cholesterol (median mg/dL, IQR) | 42372 | 166 (50) | 181 (50) | $6.17 \times 10^{-88}$ |
| HDL-C (median mg/dL, IQR) | 38483 | 47 (18.5) | 53 (22) | $2.54 \times 10^{-99}$ |
| Triglyceride (mg/dL, median (IQR)) | 42170 | 120 (74) | 107 (74) | $9.11 \times 10^{-43}$ |
| Ever smoked? (N yes, %) | 32983 | 945 (52.0%) | 10855 (34.8%) | $5.26 \times 10^{-50}$ |
| **Age groups (N,%)** | | | | $2.22 \times 10^{-308}$ |
| Less than 40 | 10608 | 35 (0.3%) | 10573 (99.7%) |  |
| 40-69 | 30123 | 1411 (4.7%) | 28712 (95.3%) |  |
| Greater than or equal to 70 | 16940 | 2320 (13.7%) | 14620 (86.3%) |  |
| **Self-reported groups (N,%)** | | | | $5.33 \times 10^{-160}$ |
| African-American | 11472 | 980 (8.5%) | 10492 (91.5%) |  |
| East/Southeast Asian | 2051 | 35 (1.7%) | 2016 (98.3%) |  |
| European American | 16720 | 545 (3.3%) | 16175 (96.7%) |  |
| Hispanic/Latino | 19574 | 1833 (9.4%) | 17741 (90.6%) |  |
| Native American | 80 | 12 (15%) | 68 (85%) |  |
| South Asian | 1484 | 48 (3.2%) | 1436 (96.8%) |  |
| Other | 1907 | 101 (5.3%) | 1806 (94.7%) |  |
| Multiple selected | 2205 | 101 (4.6%) | 2104 (95.4%) |  |
| Not available | 2087 | 107 (5.1%) | 1980 (94.9%) |  |
| **Genetic ancestry groups (%)** |  |  |  | 3.63x10<sup>-100</sup> |
| African-American/African | 7191 | 634 (8.8%) | 6557 (91.2%) |  |
| Ashkenazi Jewish | 4408 | 152 (3.4%) | 4256 (96.6%) |  |
| Non-Ashkenazi Jewish European American | 5990 | 195 (3.3%) | 5795 (96.7%) |  |
| Filipino and other Southeast Asian | 614 | 13 (2.1%) | 601 (97.9%) |  |
| Dominican | 1971 | 227 (11.5%) | 1744 (88.5%) |  |
| Ecuadorian | 438 | 31 (7.1%) | 407 (92.9%) |  |
| Puerto Rican | 5343 | 608 (11.4%) | 4735 (88.6%) |  |
| Other Central and South American | 1025 | 67 (6.5%) | 958 (93.5%) |  |

### Phenotyping using electronic health records and survey data

Biological sex at birth and age was extracted from Bio*Me* questionnaire data. Electronic Health Record (EHR) data was accessed to extract relevant disease outcome and biomarker data. Individuals were designated PAD cases based on having at least one instance of the PAD International Classification of Diseases 9th Revision (ICD-9) billing code 443.9 (from 2007 to 2018) or ICD-10 code I73.9 (from 2018 to 2020) within their EHR records from the Mount Sinai health system. Controls were defined as those with no record of either ICD-9 code 443.9 or ICD-10 code I73.9 in the EHR. Type 2 diabetes mellitus (T2D) cases and controls were defined using the Northwestern University Type 2 diabetes mellitus algorithm described in (Jennifer Pacheco and Will Thompson. Northwestern University. Type 2 Diabetes Mellitus., 2012). Triglyceride (TG; mg/dL), high-density lipoprotein (HDL; mg/dL) and total cholesterol (TC; mg/dL) lab values were extracted for all encounters for each participant (2007 to 2021). Lab values with invalid entry “999999” were removed. For each biomarker, the median value per participant was calculated. Median values per biomarker were plotted separately per sex and per self-report population label. Outlier values were defined as individuals with log10 transformed median values greater than 3rd quantile + (1.5*IQR) or 1st quantile – (1.5*IQR) (Total N participants removed per biomarker: HDL = 506, TC = 562, TG = 382). Biomarker values were then converted to z-scores for downstream analysis. The same filtering and normalization steps were applied per IBD community and per sex for IBD community based enrichment analysis (Total N participants removed per biomarker: HDL = 269, TC = 296, TG = 202). Body mass index (BMI) measures were extracted from the EHR (2007 to 2022) and median value per participant was calculated with outliers removed in the same manner as described for biomarkers for both self-reported groups (N participants removed = 963) and genetically inferred sub-groups (N participants removed = 455), and converted to z-scores.

### PAD incidence across population groups

Statistical tests for enrichment of PAD across both self-reported groups and genetically inferred sub-groups were performed using a generalized linear model (GLM) in R. PAD risk was tested separately within each self-report group relative to self-report EAs. The same models were used for association testing within each genetically inferred sub-group, this time testing for risk relative to non-Jewish Europeans. The three models for regression analysis were defined as, Model 1: PAD ∼ Population group + Age + Sex, Model 2: PAD ∼ Model 1 + BMI, Model 3: PAD ∼ Model 2 + T2D + TG + TC + HDL.

### Global ancestry inference

OMNI and MEGA genotype data was used to calculate global ancestry proportions (see **supplemental Table 1**). Reference panels of 100 individuals each representing three continental populations, African (AFR), European (EUR), and Native American (NAT), were constructed using genotype array data from the 1000 Genomes Project (1KGP), Human Diversity Genome Project (HDGP), and the Polygenic Architecture using Genetics and Epidemiology (PAGE) Study (see **supplemental Figure 2**). We randomly selected unrelated individuals from two European ancestry and two African ancestry reference populations in the 1KGP; Utah residents with Northern and Western European ancestry (CEU; N=50), Iberian populations in Spain (IBS; N=50), Yoruba in Ibadan, Nigeria (YRI; N=50), and Luhya in Webuye, Kenya (LWK; N=50) (1000 Genomes Project Consortium et al., 2015). We selected unrelated individuals with maximal NAT genetic ancestry from four Native American ancestry reference populations; an indigenous population from Oaxaca, Mexico (N=25) in HGDP (Cann et al., 2002), and indigenous populations from Honduras (N=25), Columbia (N=25) and a Peruvian population (N=25) in PAGE (Bien et al., 2016). MEGA, OMNI genotyping data was merged with the HGDP, 1KGP and PAGE reference panels using PLINK (v1.9) leaving a total of n=395531 SNPs (Purcell et al., 2007). Dominican participants were filtered to remove one of any pair of individuals who were 2nd or 1st degree relatives. Sites were filtered to remove palindromes and a minor allele frequency (MAF) threshold of 1% was applied. Linkage disequilibrium (LD) pruning was performed using PLINK according to the parameters --indep-pairwise 50 5 0.3. Regions known to be under recent selection were removed: human leukocyte antigen region (chr6:27000000-35000000,hg37), lactase gene (chr2:135000000-137000000), an inversion on chromosome 8 (chr8:6000000-16000000), a region of extended LD on chromosome 17 (chr17:40000000-45000000), ectodysplasin A receptor gene (chr2:109000000-110000000) and T Cell Receptor Beta Variable 9 gene (chr7:142000000-142500000). Following these filtering steps n=155702 SNPs and N=2133 participants remained including N=300 reference individuals with a total genotyping rate of 99%. ADMIXTURE software was used to calculate global ancestry proportions with 5-fold cross validation with K values set to 2, 3, 4, and 5 respectively (Belbin et al., 2021). Global ancestry proportions were calculated separately for Dominican community samples that had been removed due to relatedness (N=171). The same ADMIXTURE settings and reference panels were used. A nonparametric bootstrapping approach was used to calculate confidence intervals for each global ancestry proportion using the np.boot() function from the nptest package in R.

### Local ancestry inference

Eagle v2.0 was used to phase the merged MEGA, OMNI and reference panel dataset per chromosome using default parameters (Loh et al., 2016). The same filtering steps as above were used in this step but no LD pruning or MAF filter was applied, and regions defined as under recent selection were also not removed leaving a total of n=377798 SNPs for analysis. Local ancestry (LA) calling was performed on this phased dataset using RFMix V1 software with the default parameters (Maples et al., 2013). LA depth was plotted for each SNP and sites that deviated +/- 2 SD from the median were removed along with any LA calls in the HLA region (n SNPs = 371185). The haploid AFR, EUR, and NAT calls per individual were summed to obtain a LA inference derived global proportion ancestry. These LA inference derived global proportions were then compared to the proportion of global ancestry calls from the corresponding ancestry component at K=3 in the ADMIXTURE analysis, no outliers with a discordance in ancestry proportion greater than 5% were observed (see **supplemental Figure 4**).

#### Sample and site level quality control for genomic discovery

The Dominican discovery cohort used in the GWAS analysis were genotyped using either the OMNI (N = 980) or MEGA (N = 969) arrays. Both MEGA and OMNI genotyped samples were imputed to the phase 3 1KGP reference panel using SHAPEIT2 for phasing and IMPUTE2 for imputation. MEGA imputation was carried out at the University of Washington Genetic Analysis Center and quality control filtering of sites and imputation details are previously described in Wojcik *et al. (Wojcik et al., 2019)*. Imputation of OMNI genotype data, including details of genotype data quality control prior to phasing and imputation, is described in detail in (Belbin et al., 2017). Briefly, samples with plate failures, call rates <98% or deviances in rates of heterozygosity were removed. Samples with discordance between genetic and EHR recorded sex and duplicates were also removed. Sites with call rate of <95% were filtered out along with sites significantly deviating from Hardy-Weinberg equilibrium (p<1×10^-5^), calculated within ancestry groups separately. Further imputation specific quality control steps included removal of sites that failed the –miss-hap test (p < 1×10^-8^) in PLINK and duplicated sites. The phased genotype data (n SNPs= 828109 and N samples = 11212) was imputed using IMPUTE2 in 5MB chunks using the parameters “-Ne 20000 -buffer 250 -filt_rules_l ‘ALL<0.0002’ ‘ALL>0. 9998’”.

#### Admixture mapping

A total of N=1940 (N=257 cases and N=1683 controls) participants from the Dominican community, genotyped on either OMNI (N = 959) and MEGA (N = 981) arrays, were used to perform genome-wide admixture mapping. Separate LA haplotype call sets were constructed for each of the 3 ancestral groups, and haplotypes were represented as additive vectors (i.e. 0 = non-carriers, 1= heterozygous, 2= homozygous). Each LA call set was used as predictors in iterative GLMs according to the formula PAD∼LA+Sex+Array Type (R version 3.2.0). The STEAM R package was used to calculate the admixture mapping significance threshold using the get_thresh_simstat() function with nreps = 10,000 (Grinde et al., 2019). The number of generations since admixture (g) parameter was calculated using the correlation of local ancestry between pairs of pruned loci in the RFMix output. Loci were pruned to include one SNP per RFMix window (0.2 Cm). This correlation file was used with the get_g() function to calculate g=9.729661.

#### Testing generalizability of admixture mapping in Hispanic Latino populations

Two additional cohorts of HL participants were identified to determine generalizability of admixture mapping associated loci. The first cohort was an independent dataset of self-reported HL participants in Bio*Me* (N=7571) that had not been included in the Dominican discovery cohort described above. LA was inferred as above, and a GLM in R was used with the model PAD∼LA+Sex+ArrayType using NAT tracts only. The second cohort was an independent dataset of Hispanic ancestry participants (N=3675 cases, N=29558 controls) in the Million Veteran Program (MVP) (Gaziano et al., 2016). Array genotyping and quality control were described previously (Hunter-Zinck et al., 2020). LA inference was carried out using RFMIX version 2 (Maples et al., 2013). A local ancestry reference panel was constructed from the Genome Aggregate Database (GnomAD)1KGP and HGDP call set version 3.1.2 (Karczewski et al., 2020). Non-admixed (>90% estimated ancestry) samples in EUR (N=631), AFR (N=695), and NAT (N=78) populations were included in the LA inference reference panel. To ensure suitable phase quality, reference panel samples were phased in SHAPEIT4 using the TOPMed reference panel (n=194512 haplotypes) as a phasing reference. LA inference on MVP samples was performed using three rounds of expectation maximization using RFMIX. The number of inferred NAT haplotypes was then tested additively in a GLM model using PLINK 2.0, where the model included the covariates sex, age, and principal components (PCs) 1-5 (Chang et al., 2015).

#### Genome-wide association mapping

PLINK (v1.90) was used to carry out genome-wide association testing in the Dominicans using the --logistic function in both MEGA and OMNI imputed datasets separately. A MAF threshold of 1% was applied to both MEGA and OMNI imputed data. GWAS analysis using the MEGA imputed dataset (n=14227075) consisted of N=130 cases and N=838 controls. The OMNI dataset (n=13866481) included N=126 cases and N=830 controls. Quality control steps for calculating PCs is the same as that described in the LA inference, a 1% MAF filter was applied (n=281666). PCs were calculated using the SMARTPCAv10210 software from the EIGENSOFTv5.0.1 (Price et al., 2006). Age, sex and PCs 1-5 were used as covariates. METAL was used for meta-analysis of OMNI and MEGA GWAS results (n = 11614442) (Willer et al., 2010).

#### Conditional analysis

Conditional logistic regressions were used to fine map the admixture mapping signal on chromosome 2 (215-220 Mb) using R. Only participants with both local ancestry calls and 1KGP phase 3 imputed genotype data were included (N=1808). Genotype information in a 5Mb region on chromosome 2:215,041,532-219,954,973 was used and NAT haplotypes at the significant admixture mapping peak (chr2:216,636,519) were converted in the form of an additive vector of 0 = non-carriers, 1= heterozygous, 2= homozygous. Each SNP within the significant admixture mapping signal was tested as a predictor variable along with NAT tracts, sex and array type, and PAD as the outcome in iterative GLMs. The SNP which reduced the significance of admixture mapping signal by the greatest amount was SNP rs78529201 (chr2:216,518,626). An additional regression analysis then tested NAT tracts for association with PAD with rs78529201 genotype, sex and chip as covariates. Following this, the most significant NAT haplotype signal (rs7578706, chr 2:216636519) was included as a covariate along with rs78529201 genotype, sex and array type and each SNP was iteratively tested in a GLM with PAD as the outcome in order to identify an additional tag SNP.

### Plots

All plots produced using R (version 3.4.2) and ggplot2. Karyotype plots were generated using chromomap (Anand and Rodriguez Lopez, 2022). GWAS Manhattan plots and QQ plots were made using qqman and gap R packages respectively (Zhao, 2008; Turner, 2014).

## Results

### Prevalence of PAD in the diverse BioMe biobank

We assessed prevalence of PAD and associated risk factors in the diverse Bio*Me* biobank in NYC (N=57580). Overall PAD prevalence was ∼6.5%, within the 5.8-7.2% estimate reported by (Allison et al., 2007) in a study of US adults >40 years old in the year 2000 (see **Table 1**). The prevalence of PAD was lower in women (54% in cases vs 58% in controls, *P<*6.95×10^-6^) and cases were generally older (median 73 years in cases vs 59 in controls; *P<*2.22×10^-308^). PAD cases also had a slightly higher BMI (28.2 in cases vs 26.7 in controls; *P<1*.*6×10*^*-51*^) and the majority also had a T2D diagnosis (66.9% in cases vs 19.2% in controls; *P*<2.22×10^-308^). PAD cases had lower TC (166 mg/dL in cases vs 181 mg/dL in controls; *P*<6.17×10^-88^) and HDL levels (47 mg/dL in cases vs 53 mg/dL in controls; *P*<2.54×10^-99^), but higher triglyceride levels (120 mg/dL in cases vs 107 mg/dL in controls; *P*<9.11×10^-43^). A greater proportion of PAD cases had ever smoked (52% in cases vs 34.8% in controls; *P<5*.*2610*^*-50*^). As expected, PAD prevalence varied across age groups, ranging from 0.3% in participants under the age of 40, 4.7% in participants aged between 40-69 and 13.7% in participants over the age of 70 (*P<*2.22×10^-308^) (Allison et al., 2007).

We next assessed the prevalence of PAD across nine self-reported groups and eight genetic ancestry sub-groups in Bio*Me*. Genetic ancestry sub-groups were determined by community membership based on unsupervised clustering of pairwise shared IBD haplotypes that represents recent common ancestry as described in (Belbin et al., 2021). PAD prevalence was significantly different across both self-reported groups (*P<*5.33×10^-160^) and genetically inferred sub-groups (*P*<3.63×10^-100^). Within self-reported groups we observe NAs as having the highest proportion of PAD cases (15%), although sample sizes are small. The group with the second highest PAD prevalence was HLs (9.4%), followed closely by AAs (8.5%). EAsn had the lowest PAD prevalence (1.7%) consistent with reporting elsewhere (Hackler et al., 2021). Within genetically defined sub-groups, a similar pattern emerged, we observed the highest PAD prevalence in Dominican (11.5%) and Puerto Rican (11.4%) sub-groups followed by AAs (8.8%), similar to what we observed in Belbin *et al* (Belbin et al., 2021).

In order to evaluate which factors could be driving differences in observed PAD prevalence across population groups, we performed a PAD enrichment analysis. We evaluated known PAD risk factors, including age, biological sex, BMI, T2D, and lipid levels HDL-C, TG, and TC. Smoking history is another well known risk factor for PAD, however due to the high degree of missing data, it was not included in this multivariate analysis. We performed an enrichment analysis by comparing PAD prevalence in non-European ancestry to European self-reported groups (see **supplemental figure 1 and supplemental table 2)** and in genetic ancestry sub-groups (see **Figure 1 and supplemental table 3)**. In an approach adopted from Allison et al (Allison et al., 2015) we assessed three models to test for PAD enrichment, where each model added covariates adjusting for common PAD risk factors. Model 1 adjusted for age and sex, Model 2 adjusted for Model 1 and BMI and Model 3 adjusted for Model 2 and T2D, TG, TC, and HDL.

**Figure 1:**
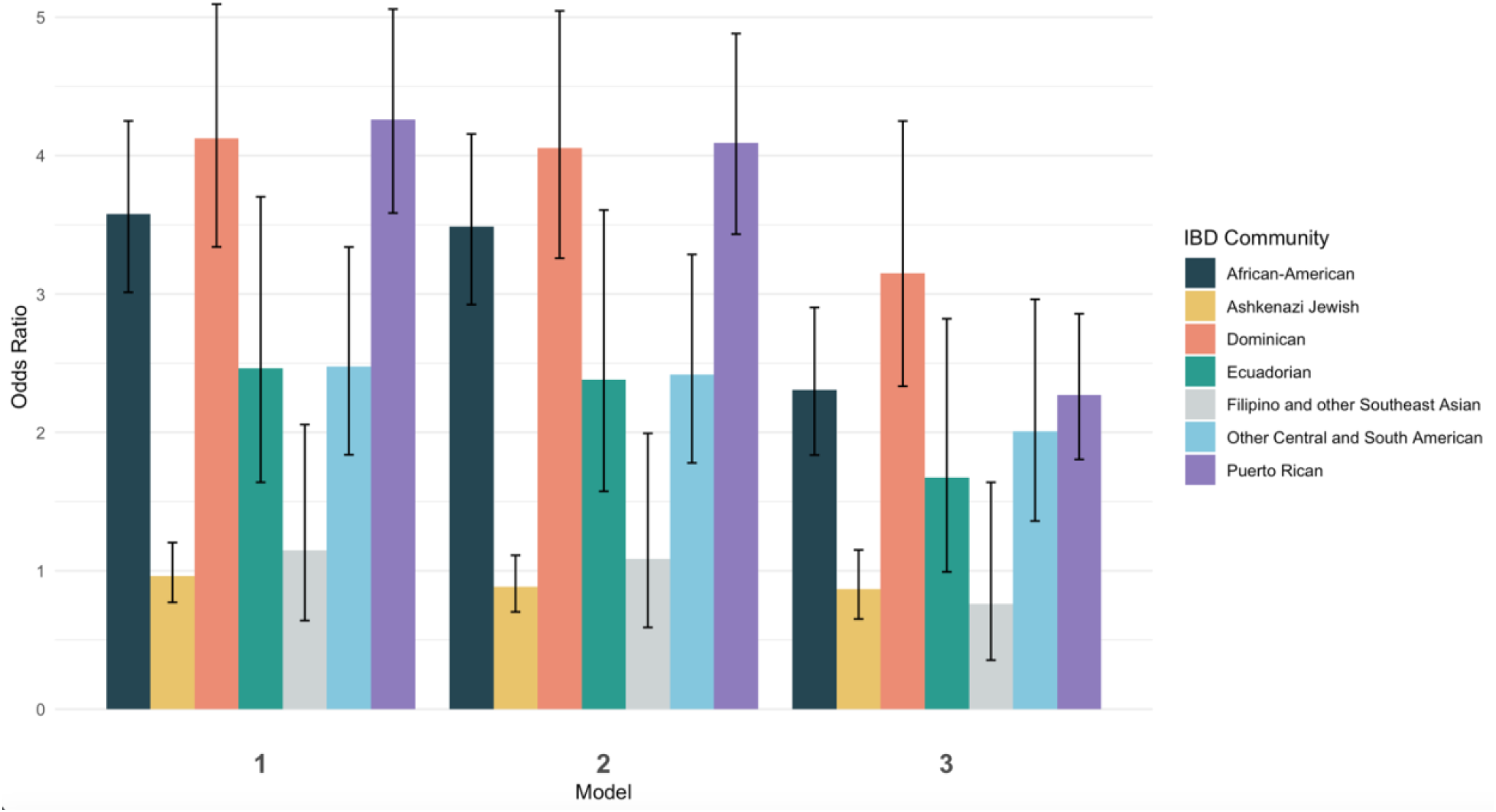
Odds of peripheral artery disease across diverse genetic ancestry groups in BioMe compared to the non-Jewish European population. Model 1: PAD ∼ Population group + Age + Sex, Model 2: PAD ∼ Model 1 + BMI, Model 3: PAD ∼ Model 2 + T2D + TG + TC + HDL.

All self-report populations are significantly enriched for PAD compared to the EA group across all models with the exception of EAsn. NAs had the highest odds ratio (OR) for PAD enrichment for all three models, and the OR in the model that included all covariates (model 3) was 6.14 (95% CI 2.75-13.69) with a P<9.33×10^-06^. However the 95% confidence interval of the OR overlaps with that observed for both HLs and AAs, and so, is not statistically different from these groups, likely due to the small sample size of NAs in Bio*Me*. Within IBD communities, all groups tested except “Filipino and other Southeast Asian” and “Ashkenazi Jewish” had a significant enrichment of PAD cases relative to the “Non-Jewish Europeans”. In Model 3 when biomarkers and T2D status is included we observed that Dominicans had highest residual increase in PAD risk (OR = 3.15 (95% CI 2.33-4.25); P= 6.44×10^-14^) compared to any other genetic ancestry sub-group in Bio*Me* (see **Figure 1**). We hypothesized that genetic risk factors common in Dominicans may be contributing to this increased PAD risk. Therefore, we decided to perform admixture mapping, which is optimally powered for genomic discovery when disease risk and/or disease-associated variants are enriched in a population.

### Global and local genetic ancestry inference in Dominicans

The first step of admixture mapping is accurate detection of genetic ancestry. First, we estimated individual-level or global genetic ancestry using ADMIXTURE, a model-based approach that applies a pre-set number of putative ancestral populations to seek the best fit of ancestral clusters in the data. ADMIXTURE analysis that was fit to three ancestral populations (k=3) recapitulates three continental-level ancestral components corresponding to AFR, EUR, and NAT reference panels (see **Supplemental Figure 2**). The median EUR and AFR genetic ancestry in the Dominican community was 56% (95% Confidence Intervals (95% CI) 55%-57%) and 37% (95% CI 36%-38%) at a population level, respectively. The majority of individuals harbored a median of 6% (95% CI 6.2%-6.5%) NAT genetic ancestry; however a small minority of individuals N=126 harbored greater than 10% (see **Figure 2A**). These global ancestry patterns were recapitulated in a principal component analysis using the same reference panels, where the majority of individuals reside on a continuum of genetic ancestry between EUR and AFR reference panels, and a small minority show appreciable NAT genetic ancestry (see **Supplemental Figure 3**).

**Figure 2:**
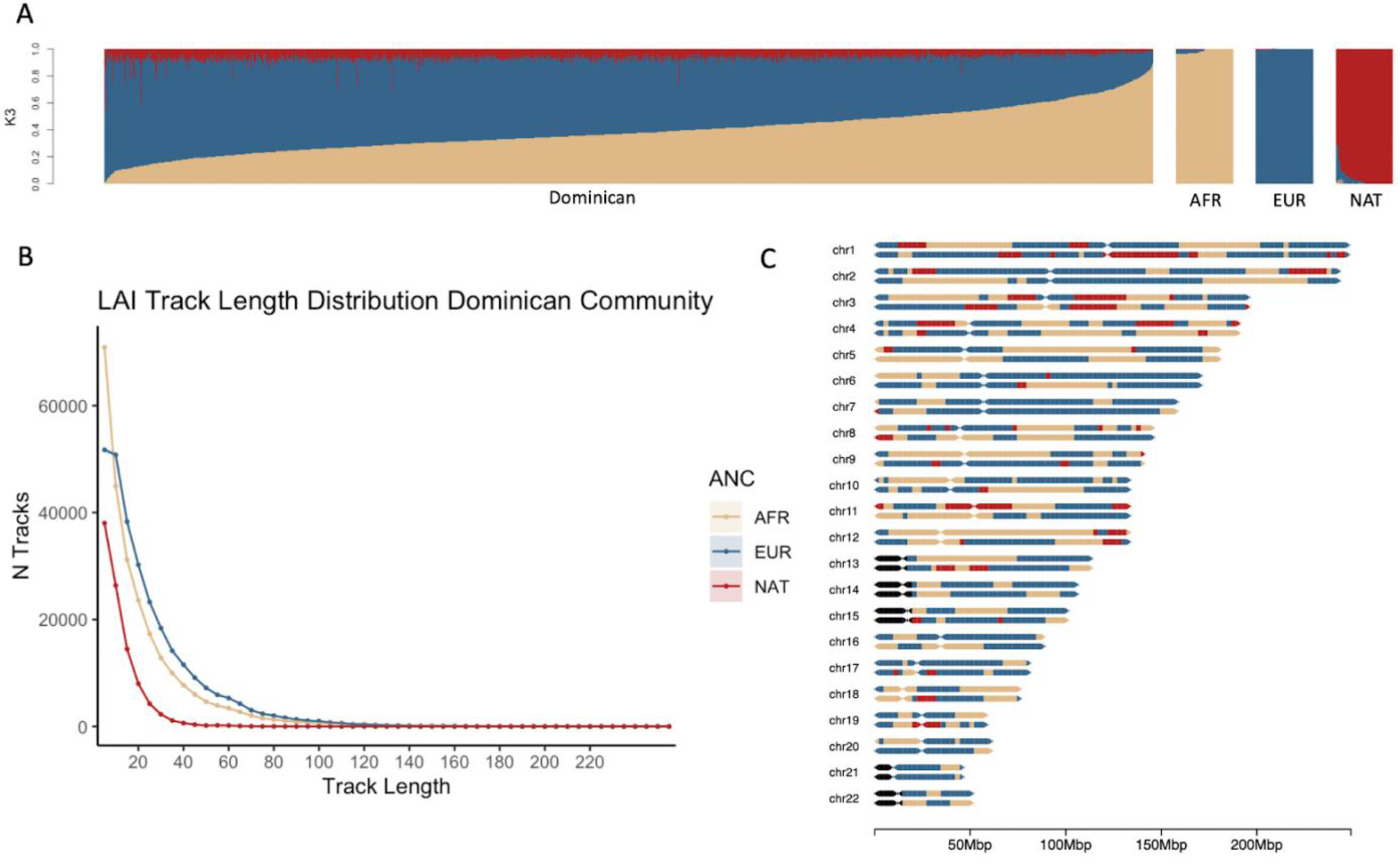
A) Admixture plots of individuals that are members of the Dominican IBD community in the BioMe biobank. Thousand Genomes Project, Human Genome Diversity Project and PAGE NAT samples were used to generate the reference panels provided. B) Tract length distribution of local ancestry haplotypes C) Karyogram plot with colors representing local ancestry tracts for a Dominican individual with approximately 6% EUR, 37% AFR and 6% NAT genetic ancestry.

To estimate haplotype-level genetic ancestry, or LA, we used RFMix, a discriminative approach that estimates tracts of LA using conditional random fields parameterized with random forests (Maples et al., 2013). We first phased the genotype data using Eagle v2.0, then ran RFMix leveraging the same AFR, EUR, and NAT reference panels as used above. Concordance between global ancestry estimates inferred using ADMIXTURE at K=3 and local ancestry estimates inferred by RFMix was high for all three ancestral components (pearson’s correlation >97%, see **Supplemental Figure 4**). **Figure 1C** shows a karyogram painted with local ancestry tracts for one individual with ∼56% EUR, 37% AFR, and 6% NAT genetic ancestry, however patterns of LAI can differ substantially between Dominican individuals. **Figure 1B** shows a plot of the tract length distribution for AFR, EUR and NAT tracts. Previous work to estimate admixture timing has suggested a single pulse of NAT ancestry contributing to Dominican genetic ancestry that occurred at the time of European contact (Moreno-Estrada et al., 2013). This is consistent with what is seen in our analysis with NAT tracts being older and shorter with a median track length of 6.6cM. EUR ancestry tracks were the longest on average (15.35 cM) followed by African tracks (11.31 cM).

### Admixture mapping of Peripheral Artery Disease in Dominicans

We next performed case-control admixture mapping to test whether regions of EUR, AFR, or NAT ancestry are associated with PAD risk. Cases for PAD (N=257) were defined as having one or more billing codes for PAD (ICD-9 443.9), and controls (N=1683) were defined as having no PAD billing codes. Admixture mapping was performed using a GLM for each ancestry group separately, including sex and array type as covariates. To estimate genome-wide significance threshold, we needed to account for the long-range correlation in local-ancestry linkage disequilibrium across the genome in the admixed Dominican community. We used the STEAM algorithm to estimate admixture proportions, generations since admixture, and genetic distances between loci in order to calculate the asymptotic joint distribution of test statistic (Grinde et al., 2019). We estimated the genome wide significance for admixture mapping in this population to be 5.282×10^-6^. A single GWS association between PAD and NAT ancestry was observed at the chromosome 2q35 locus (chr2:216636519-216811790, build 37; OR=2.05, 95% CI 1.51-2.78, P<4.06×10^-6^; see **Figure 3**). To determine whether this association might have been detected using a traditional GWAS approach, we performed GWAS in the same participants, but no GWS association was found (see **Supplemental Figure 5 for Manhattan plots**, and **Supplemental Figure 6** for QQ plot).

**Figure 3:**
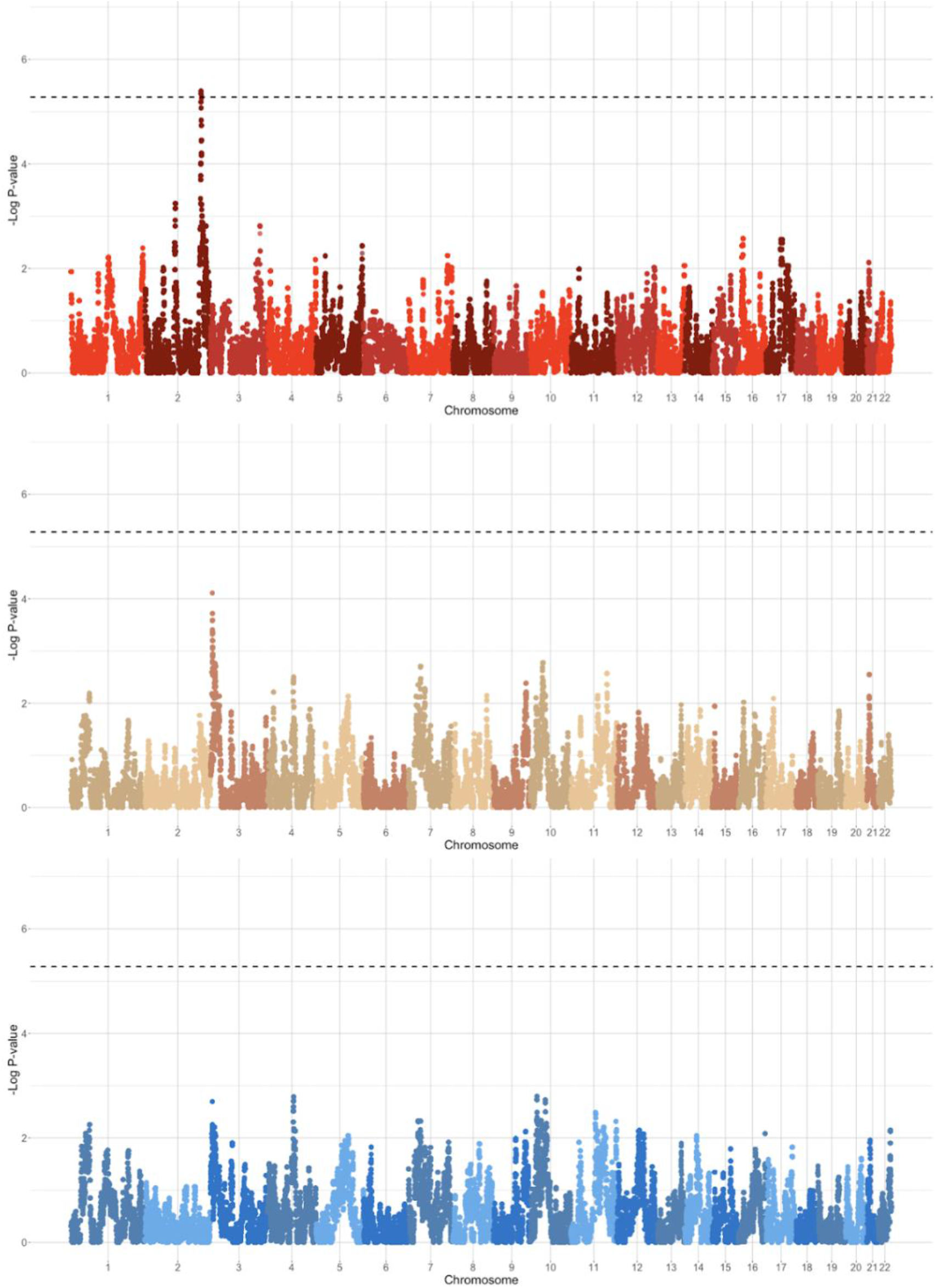
Admixture Mapping Results stratified based on EUR (Blue), AFR (Brown) and NAT (Red) haplotypes. Significance threshold after correcting for multiple testing is indicated by the dashed black line.

**Figure 4:**
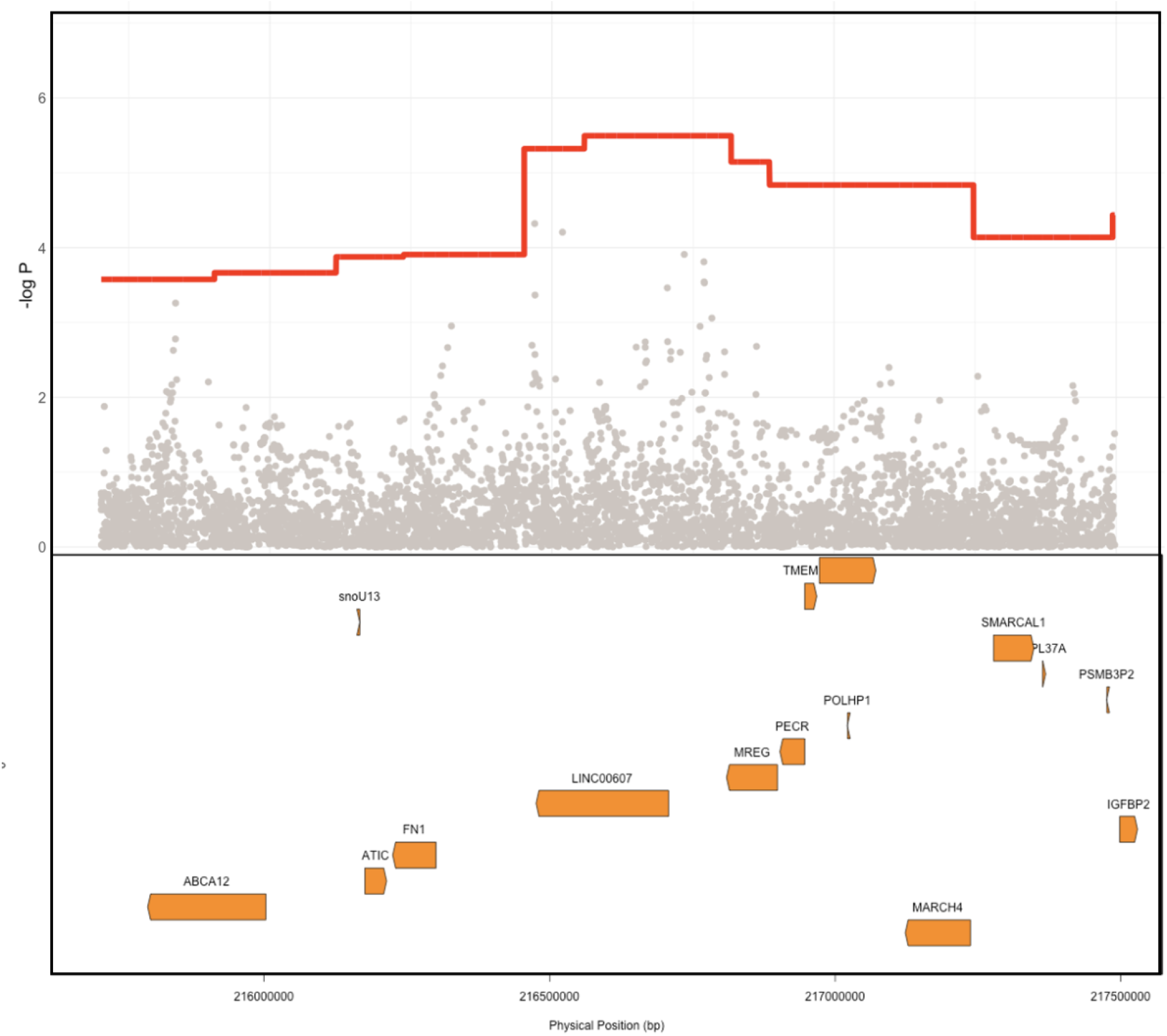
Comparison of Ancestry Signal Vs SNP level signal. NAT haplotype association is depicted in red showing a clear significant association signal compared to the individual SNP associations with PAD as the outcome. Location of genes within this haplotype are located in the lower box. Physical positions are build GRCh37.

### Characterizing the 2q35 PAD-associated locus in Hispanic Latino populations

We were unable to obtain an independent Dominican cohort with a sample size sufficiently powered to attempt replication of the 2q35 PAD-associated locus discovered in Bio*Me*. To test if the admixture mapping signal replicated broadly across HL groups, we repeated the same admixture mapping approach in self-report HLs Bio*Me* participants (N=7571), and excluded the Dominican sub-group used in the discovery analysis. The top admixture mapping signal in the 2q35 region was significantly associated, however in the opposite direction of effect, with NAT ancestry in the HL cohort being protective against PAD (OR=0.7, 95% CI 0.62-0.8, *P*<1.02×10^-07^). This suggests that the 2q35 PAD-associated locus may have population-specific effects in different HL groups. We also tested for association of NAT tracts at the 2q35 locus in the HL participants in the MVP cohort (Hunter-Zinck et al., 2020). We found no significant admixture mapping association in this region (OR=0.99, 95% CI 0.93-1.05, *P*<0.73). It is notable that the predominant HL sub-group in the Bio*Me* independent cohort are Puerto Rican descent, whereas in MVP the predominant sub-group are Mexican descent. Therefore, it is possible that these findings support a role for the 2q35 PAD-associated locus in HL populations with ancestry from the Caribbean, and not from other parts of the Americas.

### Fine-mapping the 2q35 locus

Iterative conditional analysis within a 5Mb window in this region was used for fine-mapping to identify a causal tag SNP within the NAT haplotype. Participants in the Dominican discovery cohort with both NAT haplotypes and imputed genotype calls were included (N=1808) in the analysis. A series of GLM were carried out with PAD as the outcome variable and local ancestry status at position 2:216636519 (build 37) (0=no NAT ancestry, 1=heterozygous NAT, 2=homozygous NAT), test tag SNP, age and sex as predictor variables. Inclusion of the tag SNP in this conditional logistic regression with the top local ancestry association signal causes the ancestry haplotype significance to increase towards zero. Using this approach, we identified tag SNP rs78529201 (chr2:216518626, build 37) which attenuates the p-value of the local ancestry (from P<3.2×10^-06^ to P<0.0046; see **Figure 5A**). Though this SNP was able to explain much of the signal, it was not sufficient to fully explain the PAD-associated signal at 2q35. Therefore, the windowed conditional analysis was carried out again to find a likely second contributory tag SNP in the region. The SNP rs77979649 (chr2:218477448, build 37) upstream of the primary tag SNP when included in a model with rs78529201, the admixture mapping signal was fully attenuated (P<0.312). Neither SNP met genome wide significance using the traditional GWAS approach, rs78529201(OR=3.63, CI 95% 1.69-7.79, P<9.3×10^-04^) and rs77979649 (OR=2.14, CI 95% 1.09-4.20, P<2.7×10^-02^). Finally, we examined the frequency of both SNPs in the gnomAD browser v3.2.1 (Karczewski et al., 2020). They are both common (MAF>5%) in the Latino/Admixed American population, and rs77979649 is also common in the East Asian population, and low frequency (MAF<5%) or absent in other populations in the gnomAD. In the gnomAD Latino/Admixed American group, they are at high frequency (MAF>25%) on Amerindigenous LA tracts and rare (MAF<0.1%) on AFR and EUR LA tracts. This suggests that both tag SNPs are highly differentiated on NAT tracts in Bio*Me* Dominicans and the Amerindigenous LA tracts in the gnomAD Latino/Admixed Americans population, however neither SNP perfectly tags the NAT signal at 2q35 in Bio*Me* Dominicans.

**Figure 5:**
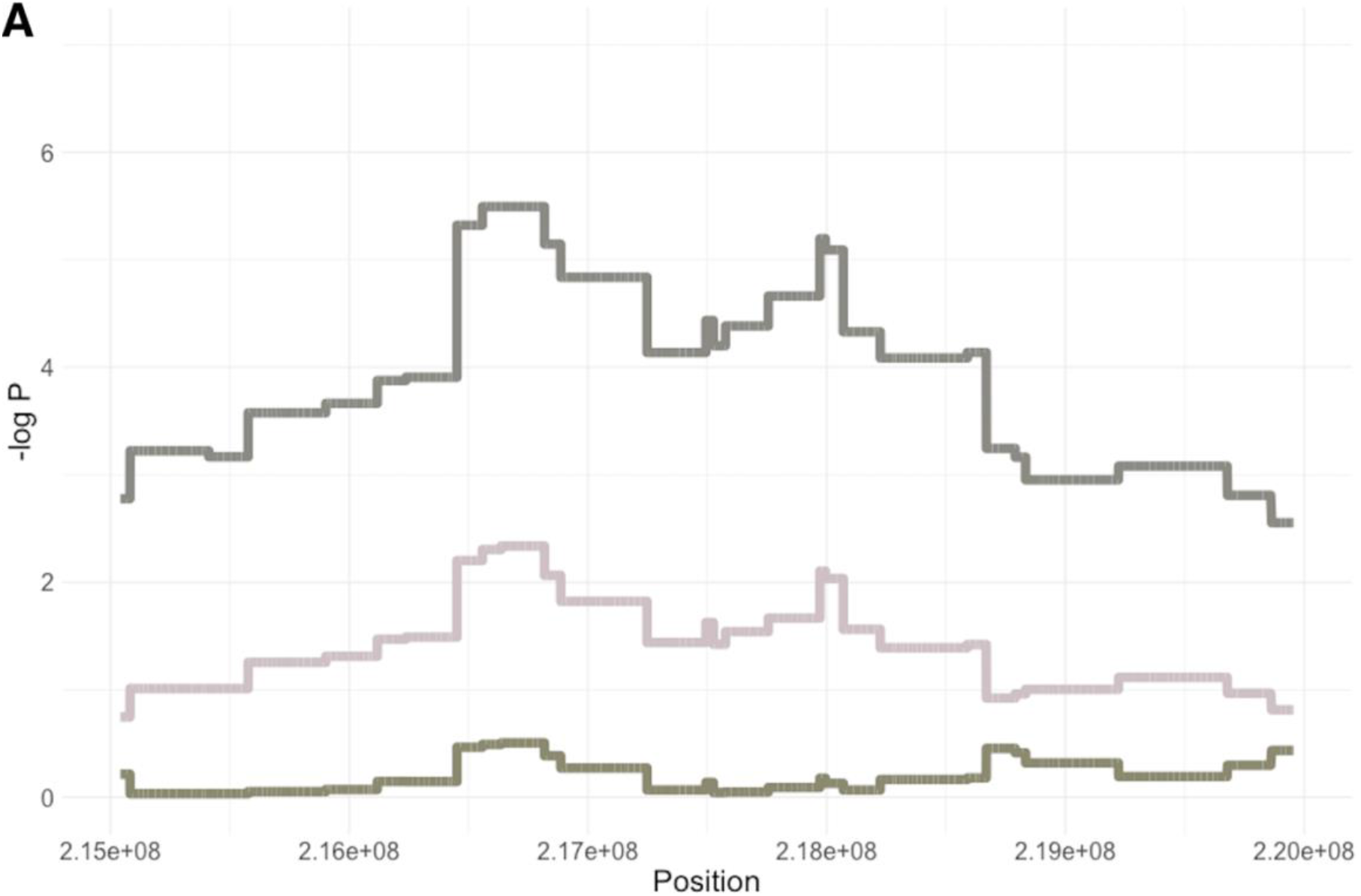
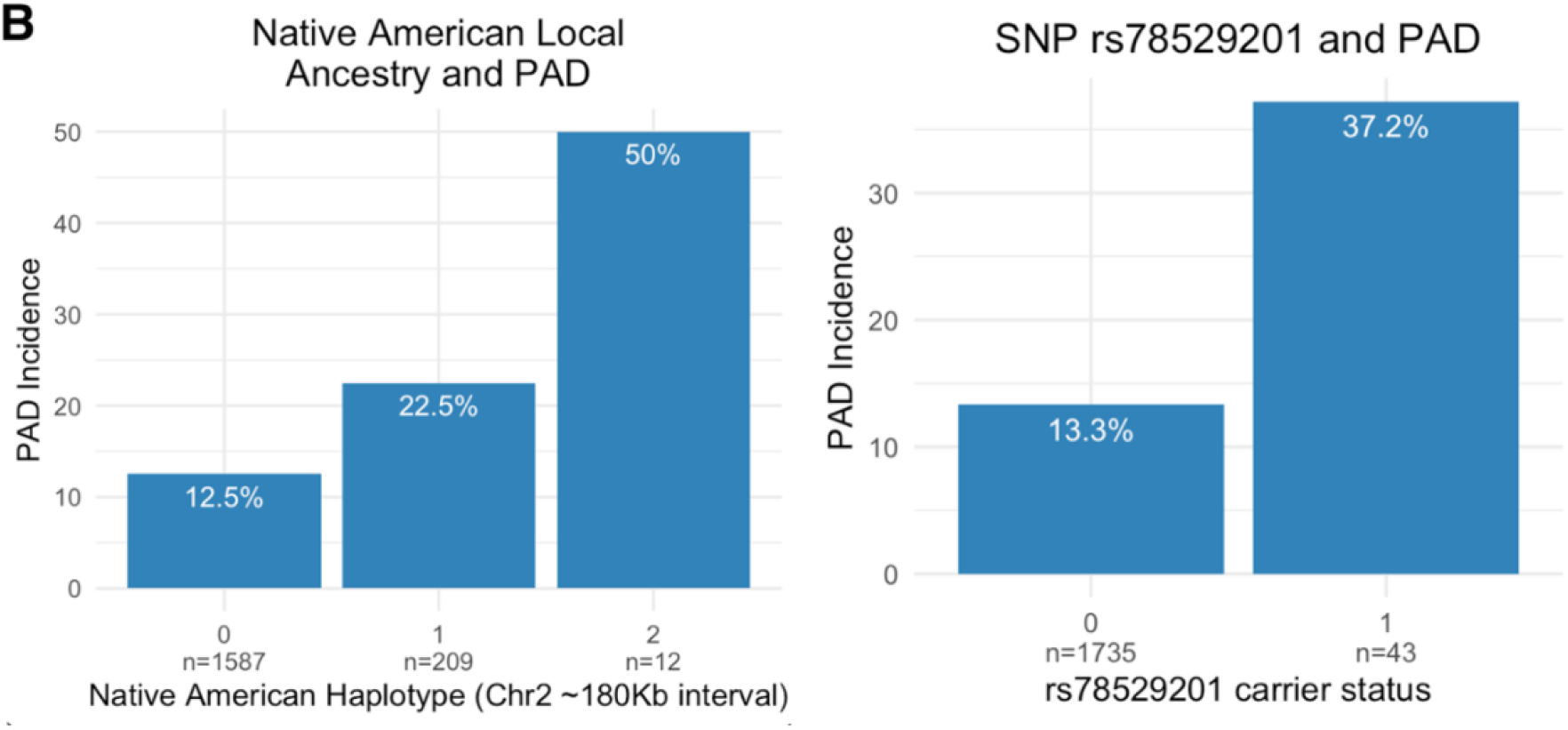
**A)** Fine-mapping the 5Mb region on chromosome 2 (215,000,000-220,000,000) using conditional analysis. Two tag SNPs, rs78529201 and rs77979649, increase the NAT admixture mapping signal towards 0 when included in the association model **B)** Breakdown of PAD incidence by NAT haplotype and rs78529201 carrier status (n = sample size).

The top tag SNP, rs78529201, falls within an intron of the lincRNA, *LINC00607*. Bulk tissue gene expression results from the Genotype-Tissue Expression portal (GTEx Portal) show the tissue with the highest expression of *LINC00607* are arteries (Sriram et al., 2022) and another recent study demonstrated it is highly enriched in endothelial cells (Boos et al., 2022). Previous work has shown a key role of *LINC00607* in the control of cellular processes underlying inflammatory responses, angiogenesis, collagen catabolism and extracellular matrix organization in endothelial cells via its inactivation of *PAI-1*, an inhibitor of plasminogen activators (Calandrelli et al., 2019; Sriram et al., 2022). To assess the impact of harboring a risk allele on PAD risk, we showed that Dominican individuals who are heterozygous for SNP rs78529201 show a 37.2% risk of the disease compared to 13.3% of non-carriers (OR=3.88, CI 95% 2.06-7.31, *P*<1.95×10^-5^). No homozygous carriers are present in the Bio*Me* biobank for this variant. We also show that heterozygous carriers of a NAT haplotype in the 2q35 region carry a 22.5% risk of PAD, compared to 12.6% for non-carriers (OR=2.02, 95% CI 1.42-2.89, *P*<0.00013) (see **Figure 5B**). There are only 12 homozygous carriers of a NAT haplotype at rs76984916, prohibiting statistical comparison with non-carriers.

## Discussion

In this study we assessed clinical, demographic, and genetic factors underlying PAD risk in the diverse Bio*Me* biobank in NYC. Among genetic ancestry defined sub-groups, Dominicans had the highest enrichment of PAD (3-fold compared to European ancestry Bio*Me* participants), when accounting for known clinical and demographic PAD risk factors. Local ancestry inference delineated haplotypes of recent AFR, EUR, and NAT continental ancestry in the Dominican group, and admixture mapping revealed a strong signal of association of NAT ancestry tracts on chromosome 2q35 linked to a 2-fold increase in PAD risk. Individuals who are heterozygous for NAT at the 2q35 locus have 22.5% incidence of PAD in Bio*Me* compared to 12.6% in those with no NAT at that locus. Fine-mapping revealed a top tag SNP, rs78529201, which falls within an intron of the lincRNA, *LINC00607. LINC00607* has been previously shown to play a key role in the control of cellular processes underlying angiogenesis, extracellular matrix organization, and other vascular-related processes, in endothelial cells. These findings highlight a previously under-appreciated risk for PAD, and a novel genetic driver of increased PAD risk at chromosome 2q35, in Dominican populations in NYC.

Within self-reported populations in our study, we found that HLs and AAs had similar enrichment (∼2.6-fold) of PAD compared to EAs. This finding differs from some previous studies which have reported AAs as having the greatest PAD risk among race and/or ethnicity groups in the US (Allison et al., 2006). HLs have been reported to have lower or similar rates of PAD compared to EAs, despite having a paradoxically high burden of PAD risk factors, however the picture appears to be nuanced (Criqui et al., 2005; Forbang et al., 2014; Allison et al., 2015). Allison et. al. reported Puerto Ricans and Dominicans have higher rates of PAD relative to Mexican Americans. Using genetic ancestry to identify HL sub-groups in NYC, we were able to explore incidence of PAD in four sub-groups; three were significantly enriched compared to the European ancestry population; Puerto Ricans (2.3-fold), Dominicans (3.2-fold), and Central and South Americans groups (2-fold). Factors not measured in this study, including structural racism, socioeconomic status, disparities in healthcare access, detailed smoking habits, statin use, discussed further by (Hackler et al., 2021), are all likely contributing to differences in PAD incidence both at local and national-levels. But this study, along with work from the Hispanic Community Health Study/Study of Hispanics (Lavange et al., 2010) and Multi-Ethnic Study of Atherosclerosis (Bild et al., 2002), demonstrates that HLs are culturally, socioeconomically, and genetically heterogeneous, and disease incidence rates may differ across HL sub-groups. We note that, although a small sample size, the self-reported NA group in Bio*Me* had the highest PAD enrichment (6-fold), warranting further study of PAD risk factors in this population.

Admixture mapping of PAD in a Dominican population from NYC identified a single significantly associated region on chromosome 2q35. The risk locus spanned a 0.2 MB region on chromosome 2, where NAT genetic ancestry was associated with increased risk for PAD. An association with NAT genetic ancestry at the same locus, but in the opposite direction was demonstrated in the same biobank in an independent cohort of self-report HLs of predominantly Puerto Rican descent, and no association was observed in the HLs in the MVP, who are predominantly of Mexican descent (Gaziano et al., 2016). It is possible that the observed differences in PAD-association at the 2q35 locus may be tied to patterns in NAT population structure in admixed populations from the Americas. Previous work has demonstrated population-substructure in the NAT ancestry components in HL populations with origins from Mexico (Moreno-Estrada et al., 2014) and Caribbean (Moreno-Estrada et al., 2013), and genetic divergence between ancestral NAT populations (Reich et al., 2012). The interaction of the 2q35 with social or environmental factors impacting PAD may also modulate its impact across HL groups.

Fine-mapping of the associated locus revealed a top tag SNP residing in the intronic region of a lincRNA *LINC00607*. LincRNAs are known to play an important role in gene expression, usually in a cell type specific manner, and have recently been shown to play an important role in complex disease (Deniz and Erman, 2017). Recent work to characterize *LINC00607* expression and gene regulatory networks in vascular smooth muscle cells and endothelial cells, found it to be an essential regulator of vascular cell function. Temporal changes to gene expression in endothelial cells mimicking diabetic conditions induced an intrachromosomal interaction between *LINC00607* and a super enhancer overlapping the SERPINE1/PAI-1 gene (Calandrelli et al., 2020). *LINC00607* knockout reduced *SERPINE1* expression along with numerous other genes including *FN1, TRIO* and *COL4* via a super enhancer network, promoting endothelial cell dysfunction (Calandrelli et al., 2019). *LINC00607* is also implicated as a vital epigenetic regulator in arterial tissue and potential target for CVD therapies (Sriram et al., 2022). Taken together, this supports a role for *LINC00607* as a gene expression regulator of key genes related to extracellular matrix organization, inflammation, angiogenesis and endotheliopathy, suggesting a putative link of the 2q35 locus to PAD etiology.

There are several limitations to this study. First, cohorts with origins from the Dominican Republic are vastly underrepresented in genomic research databases (Estrada-Veras et al., 2016), and we couldn’t access an independent cohort of Dominican ancestry of sufficient size and with the relevant phenotype to replicate our finding. Without replication, we cannot rule out association due to biases (Kraft et al., 2009) or effect size estimate confounding due to winners curse (Zou et al., 2022). Additional genetic studies of PAD in Dominican populations are needed to confirm association, examine differences in LD and effect sizes, and assess any effect modifiers across studies. Second, deriving phenotype information from health systems data can result in selection bias and confounding that can affect case ascertainment and prevalence estimates (Haneuse and Daniels, 2016), although we note that the prevalence estimates reported here are in line with previously published estimates from large cohort studies. Third, we were not able to assess the impact of some known risk factors for PAD, such as smoking. Previous studies have demonstrated the odds ratio for symptomatic PAD in smokers is 2.3 (Willigendael et al., 2004). Ideally smoking status would have been included as a covariate in our models, however smoking is not well captured in Bio*Me* questionnaire responses with >40% missing data, and was excluded from the multivariate analysis due to low power. Finally, small sample sizes for individuals self-identifying as NA limited our ability to estimate prevalence and PAD enrichment in this group with strong confidence. Ongoing efforts, to enhance ethical genomic research with Indigenous communities (Claw et al., 2018), studies focused on underrepresented populations like Human Heredity and Health in Africa (H3Africa Consortium et al., 2014), and large biobank initiatives that enrich for admixed and diverse populations, such as Bio*Me (Belbin et al., 2021)* and the All of Us Research Project (All of Us Research Program Investigators et al., 2019), will provide data that may improve power for discovery efforts in the future.

This work demonstrates how genomic discovery pipelines that leverage recent patterns of demography can be better powered to elucidate disease risk variants over traditional approaches. Furthermore, extensive population diversity often encountered in urban healthcare systems, offers the potential to examine the generalizability of risk variants across populations, the interplay with clinical and social determinants of health, and to identify populations at higher risk due to these factors. This is particularly important for complex conditions that are historically underdiagnosed. In the case of PAD, only 10% to 30% of patients show intermittent claudication which is an important sequela of PAD. The Peripheral Arterial Disease Awareness Risk and Treatment: New Resources for Survival study found that only 45% of a cohort with PAD had been diagnosed (Hirsch et al., 2001). Therefore, a better understanding of the genetics of the disease in diverse populations could help identify groups at higher risk for follow up care and prevention and ensure equity and implementation of precision medicine globally.

## Data Availability

All data produced in the present study are available upon reasonable request to the authors.

## Acknowledgements

We are grateful to the patients and their families who contributed to this study.

## Abbreviations

PAD: Peripheral artery disease
AA: African American
EA: European American
HL: Hispanic/Latino
NYC: New York City
US: United States
LA: Local ancestry
EUR: European
AFR: African
NAT: Native American
GWAS: Genome Wide Association Study
MVP: Million Veteran Program
GWS: Genome Wide Significant
SNP: Single nucleotide polymorphism
lincRNA: Long intergenic non-coding RNA
EAsn: East/South-East Asian
SA: South Asian
NA: Native American
OMNI: Omni-express array
MEGA: Multi-ethnic genotype array
IBD: identical by descent
PPV: positive predictive value
EHR: Electronic Health Record
ICD: International Classification of Diseases
T2D: Type 2 Diabetes
TG: Triglyceride
HDL: High-density lipoprotein
TC: Total cholesterol
BMI: Body Mass Index
GLM: Generalized linear model
1KGP: 1000 Genomes Project
HDGP: Human Diversity Genome Project
PAGE: Polygenic Architecture using Genetics and Epidemiology
CEU: Utah residents with Northern and Western European ancestry
IBS: Iberian populations in Spain
YRI: Yoruba in Ibadan, Nigeria
LWK: Luhya in Webuye, Kenya
MAF: Minor allele frequency
LD: Linkage Disequilibrium
GnomAD: Genome Aggregate Database
PCs: Principal Components
OR: Odds Ratio

