## Supplemental Figures and Tables for "Admixture Mapping of Peripheral Artery Disease in a Dominican Population Reveals a Novel Risk Locus on 2q35"

Supplementary Figures Legends and Tables

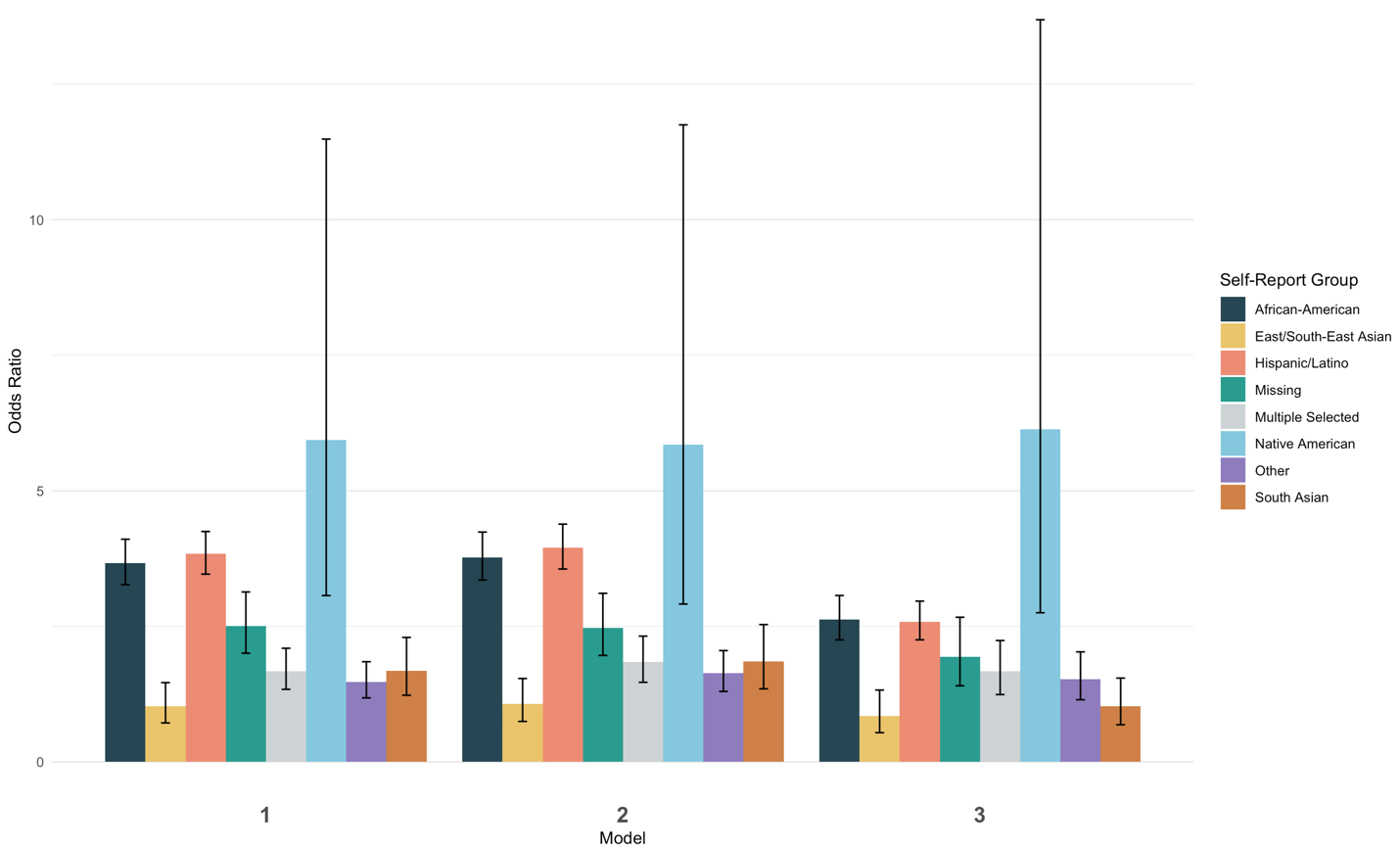

***Supplemental Figure 1 :*** *Odds of peripheral artery disease across seven self-report groups, and one group with missing data, in BioMe compared to the self-reported European population. Model 1: PAD ~ Population group + Age + Sex , Model 2: PAD ~ Model 1 + BMI, Model 3: PAD ~ Model 2 + T2D + TG  + TC + HDL.*

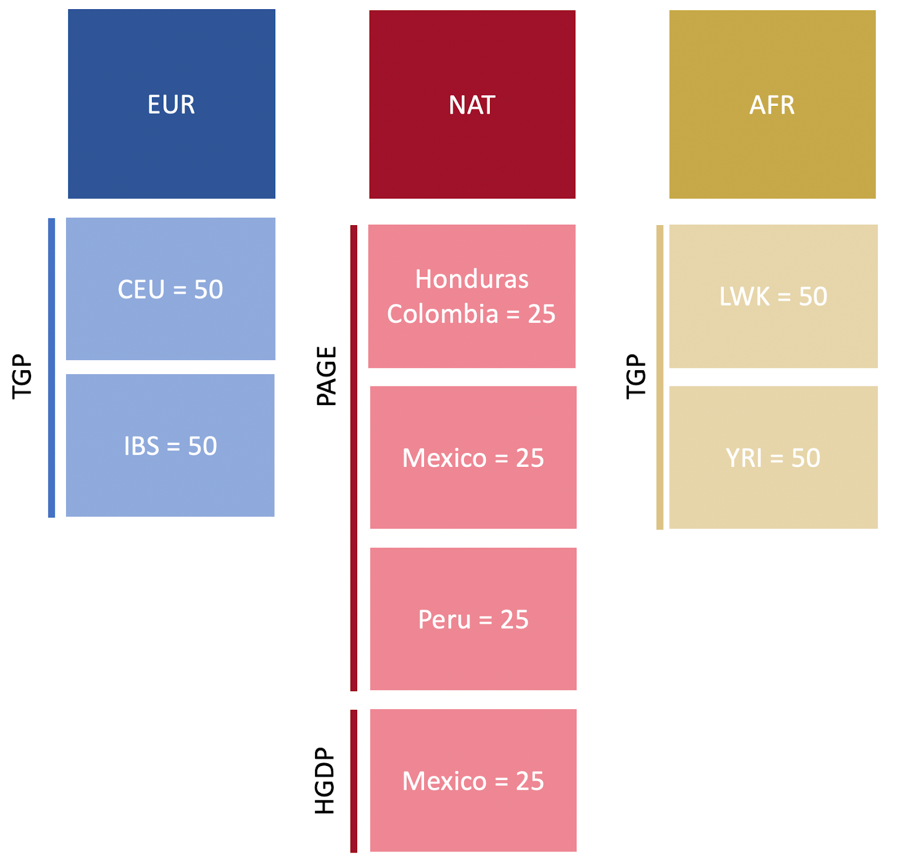

***Supplemental Figure 2:*** *Schema of reference panel used for local ancestry inference. This reference panel is also included in admixture and PCA for interpretation purposes*

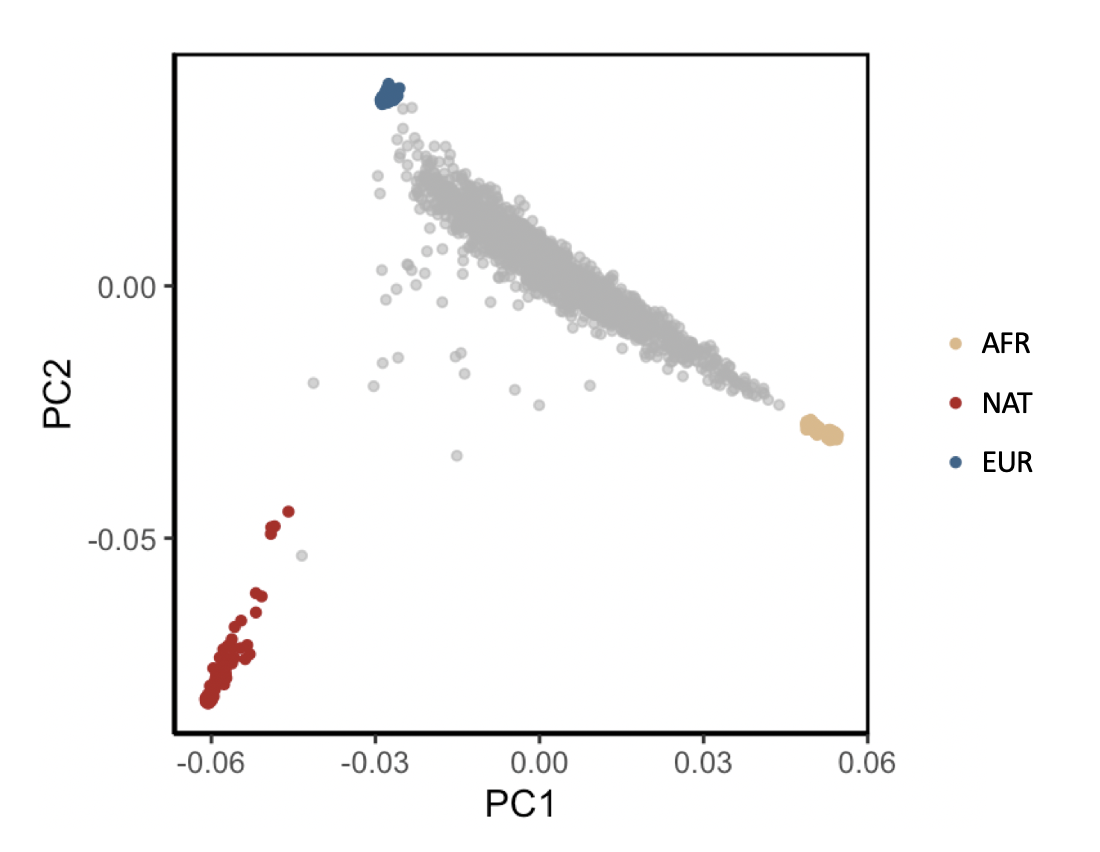

***Supplemental Figure 3:*** *PCA plot of Dominican participants included in admixture mapping.*

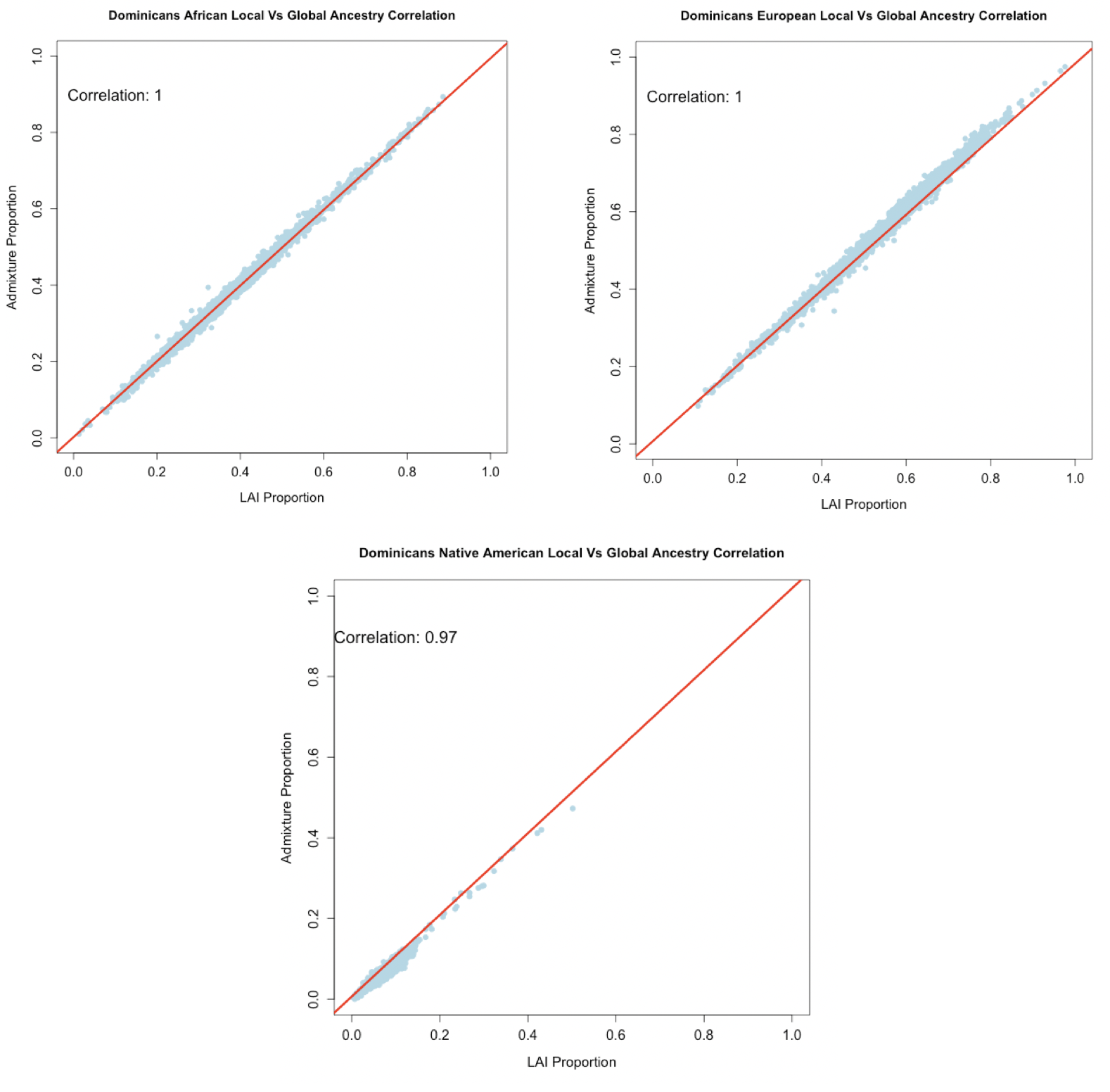

***Supplemental Figure 4:*** *Quality control of local ancestry calls. Local ancestry calls are summed per individual and compared to global ancestry proportions calculated using Admixture software.*

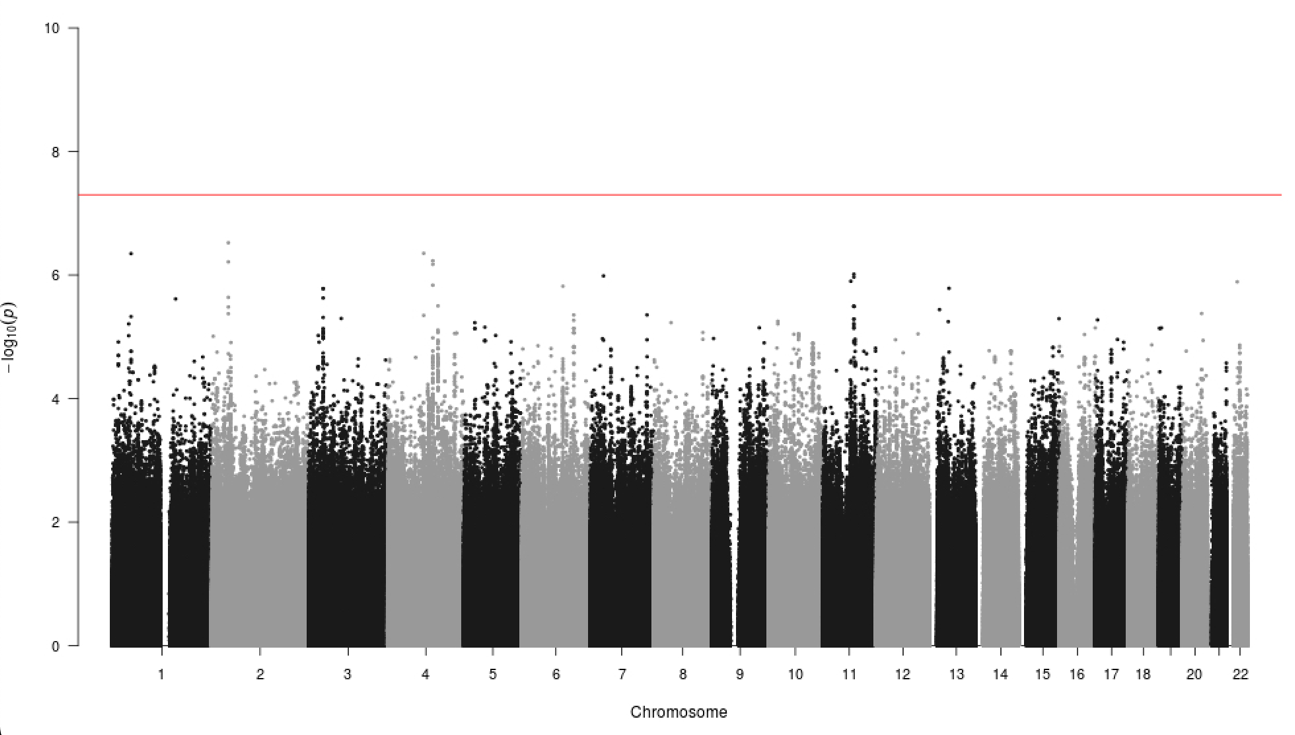

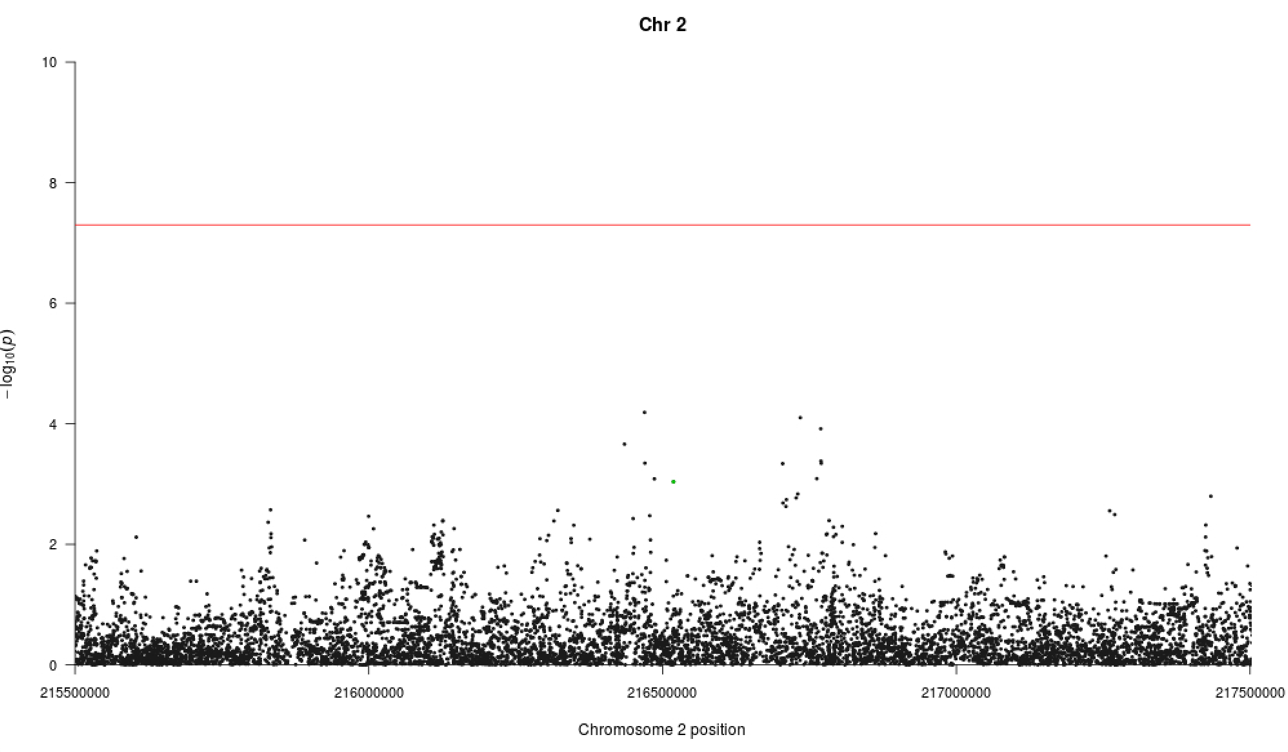
***Supplemental Figure 5: A)*** *Manhattan plot showing -log_10_ p-values from PAD GWAS meta-analysis in the BioMe Dominican discovery cohort using imputed MEGA and OMNI genotype data. Genome wide significance threshold shown in red.* ***B)*** *Zoom of Manhattan plot A highlighting* 2q35 region, tag SNP rs78529201 is shown in green.

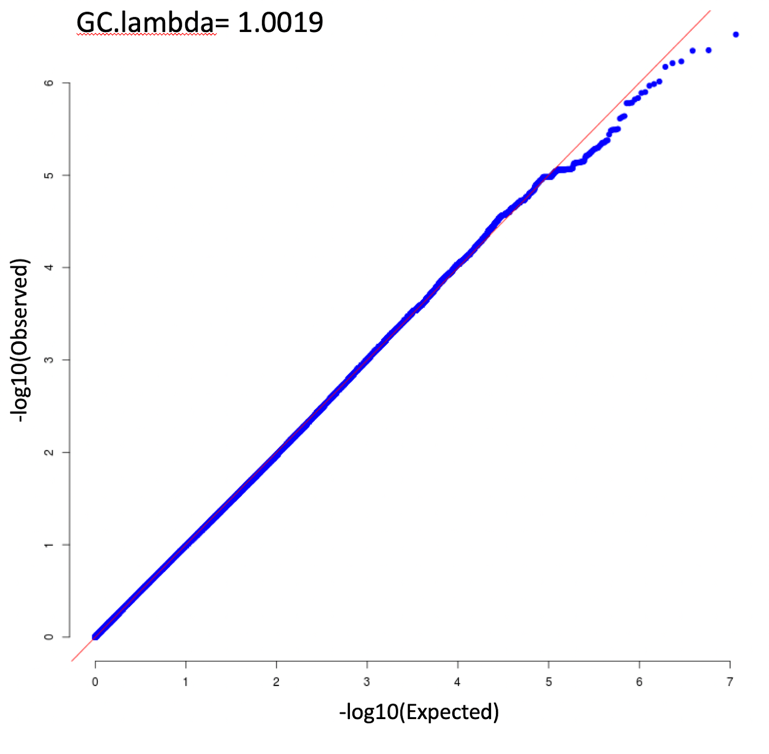

***Supplemental Figure 6:*** *QQ plot of -log_10_ p-values from PAD GWAS meta-analysis in the BioMe Dominican discovery cohort using imputed MEGA and OMNI genotype data.*

|  | Sample    Size | N    Female (%) | N PAD    Cases (%) | MedianAge    (SD) | Genotyping    Panels |
| --- | --- | --- | --- | --- | --- |
| Dominicans   (Admixture Mapping) | 1940 | 1284  (66.2%) | 257    (13.2%) | 56    (16.68) | MEGA    and OMNI |
| Dominicans   (GWAS) | 968 | 632    (65.2%) | 128    (13.2%) | 58    (14.65) | MEGA |
|  | 956 | 628    (65.7%) | 126    (13.2%) | 55    (17.16) | OMNI |

***Supplemental* Table 1:** *Summary of Dominican cohort used in discovery admixture mapping analysis and GWAS.*

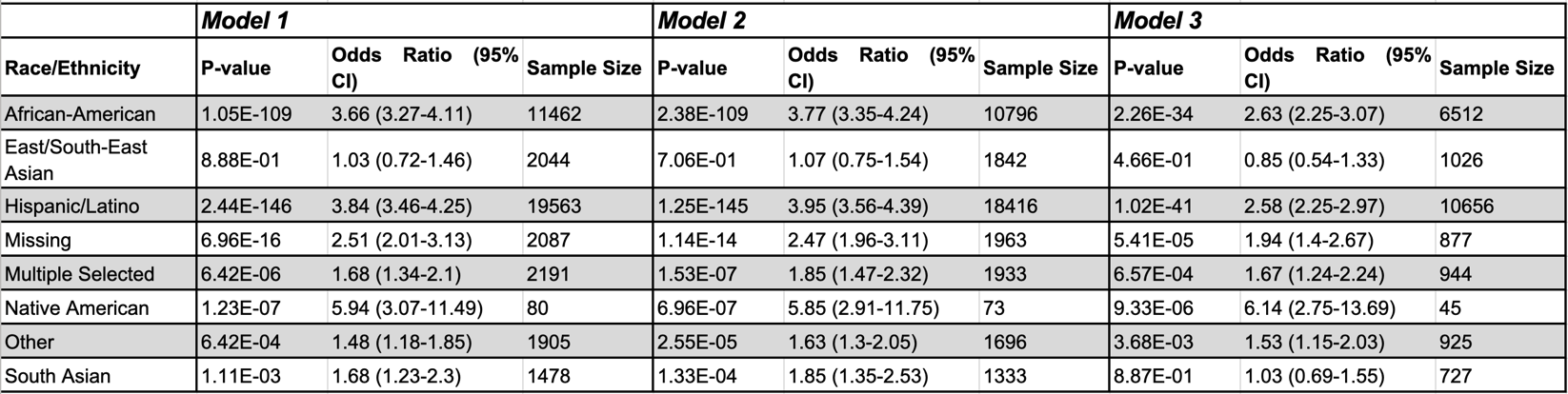

***Supplemental Table 2:*** *Results of enrichment analysis of peripheral artery disease across diverse genetic ancestry groups in BioMe compared to the self-reported European population. Model 1: PAD ~ Population group + Age + Sex, Model 2: PAD ~ Model 1 + BMI, Model 3: PAD ~ Model 2 + T2D + TG  + TC + HDL.*

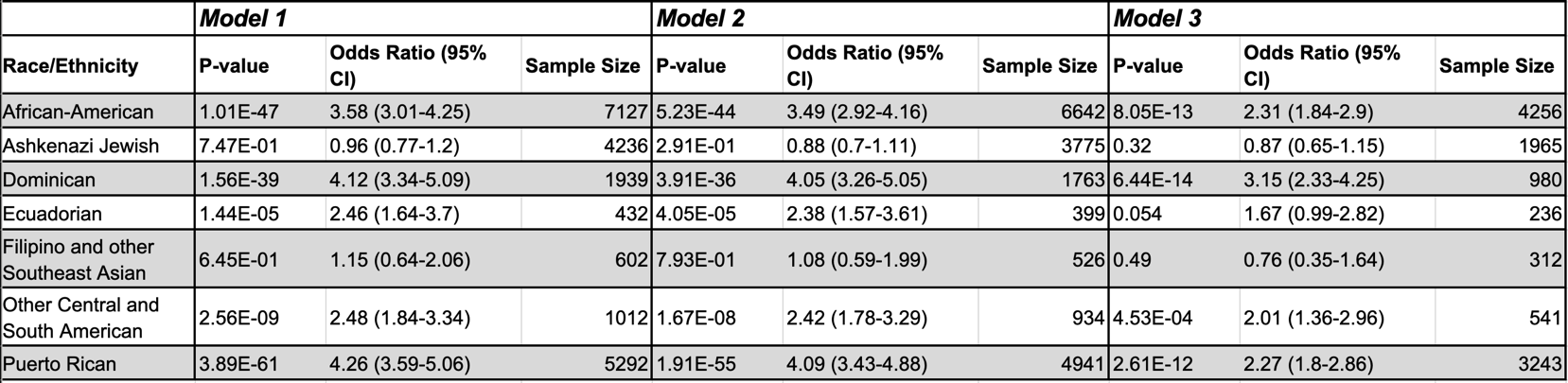

***Supplemental Table 3:*** *Results of enrichment analysis of peripheral artery disease across diverse genetic ancestry groups in BioMe compared to the non-Jewish European population. Model 1: PAD ~ Population group + Age + Sex, Model 2: PAD ~ Model 1 + BMI, Model 3: PAD ~ Model 2 + T2D + TG  + TC + HDL.*
